# Diagnostic Utility of Multi-Domain Language Measures Across the Alzheimer Disease Continuum

**DOI:** 10.64898/2026.09.28.26364143

**Authors:** Zihao Geng, Maxime Montembeault, Valérie Coulombe

## Abstract

**Objectives:** Alzheimer’s disease (AD) progresses from a cognitively unimpaired stage through mild cognitive impairment (MCI) to dementia. Because language impairment often emerges before dementia, it is a candidate marker of disease stage. Yet the standard batteries of large AD research cohorts rely on a narrow set of tests that mainly evaluate the lexical-semantic domain (i.e., picture naming and verbal fluency), leaving other language domains, such as semantic memory, phonology, morphosyntax, and single-word reading, rarely assessed. We evaluated the diagnostic utility of standard and specialized language tests for distinguishing amyloid-positive MCI and dementia of the Alzheimer’s type (DAT) from healthy controls (HC) and from each other.

**Methods:** We included 286 participants from the National Alzheimer’s Coordinating Center (125 HC, 53 MCI Aβ+, 108 DAT Aβ+) assessed with three standard tests from UDSv3 (semantic fluency, letter fluency, and Multilingual Naming Test [MINT]) and nine specialized tests from the FTLD Module (regular and irregular word reading, semantic word-picture matching, semantic associates, Northwestern Anagram Test – Short Form [NAT-S], Northwestern Naming Battery noun and verb naming, sentence repetition, and sentence reading). For each test, the area under the receiver operating characteristic curve (AUC) was computed for DAT vs. HC, MCI vs. HC, and DAT vs. MCI, and AUCs were compared between tests with DeLong’s test.

**Results:** Eight of 12 measures reached acceptable discrimination (AUC ≥ .700) for DAT vs. HC, five for MCI vs. HC, but only semantic fluency and NAT-S for DAT vs. MCI. Semantic fluency showed the highest discrimination in each contrast, though it did not significantly outperform the NAT-S for MCI vs. HC or DAT vs. MCI. MINT, letter fluency, and irregular word reading reached clinically meaningful accuracy in distinguishing MCI and DAT from HC, but not DAT from MCI. The remaining specialized tests reached it only for DAT vs. HC, or not at all. In a post hoc regression, adjusting for cognitive flexibility and working memory attenuated group differences; the DAT difference was no longer significant, whereas the MCI difference remained significant.

**Discussion:** Semantic fluency was the strongest single language marker across the AD continuum. Among specialized tests, NAT-S was the most informative, challenging the view that syntax is spared in early AD. Its performance was partly explained by executive function, mainly in DAT, whereas a residual morphosyntactic component remained in MCI, and several specialized tests were limited by ceiling effects. Clinically, the NAT-S may complement semantic fluency for staging, and specialized tests may help characterize heterogeneous language profiles in MCI and DAT.

## 1. Introduction

Alzheimer’s disease (AD) is a progressive neurodegenerative disorder that leads to cognitive decline, reduced quality of life, and increased dependence in daily activities (<u>Alzheimer’s Association, 2025</u>). Its underlying pathological processes develop gradually and can now be detected and staged using biomarkers (<u>Jack et al., 2024</u>). Under current criteria,biological AD is defined by biomarker evidence of amyloid-β pathology, while biomarker evidence of tau pathology provides additional evidence of AD and can be used to stage its biological progression (<u>Jack et al., 2024</u>). As the disease progresses, cognitive and functional changes generally unfold along a clinical continuum from a cognitively unimpaired stage to mild cognitive impairment (MCI) and, ultimately, dementia (<u>Albert et al., 2011</u>; <u>Jack et al., 2024</u>; <u>McKhann et al., 2011</u>). The typical amnestic dementia syndrome associated with AD, *dementia of the Alzheimer’s type (DAT)*, is characterized by episodic memory impairment and impairment in at least one additional cognitive domain such as executive function, visuospatial processing, or language (<u>McKhann et al., 2011</u>). This typical presentation should be distinguished from atypical, non-amnestic variants of AD, most notably the logopenic variant of primary progressive aphasia, in which language impairment is the predominant early feature (<u>Gorno-Tempini et al.,</u> <u>2011</u>; <u>Montembeault et al., 2018</u>). Throughout this study, DAT refers exclusively to the typical amnestic presentation. MCI is a clinically heterogeneous, transitional stage in which cognitive difficulties are measurable but functional independence is largely preserved. MCI can be further classified by the presence of episodic memory impairment (i.e., amnestic vs. non-amnestic) and the number of cognitive domains affected (i.e., single-domain vs. multidomain) (<u>Petersen, 2004</u>;<u>Winblad et al., 2004</u>). Sensitive clinical markers of AD-related change are nonetheless needed to distinguish stages along the AD continuum, improve early detection, monitor progression toward DAT, and deliver early intervention to prevent or delay disease progression (<u>Albert et al., 2011</u>; <u>Petersen, 2004</u>; <u>Rasmussen & Langerman, 2019</u>).

Memory impairment has traditionally dominated the diagnostic framework for DAT, but converging evidence indicates that language impairment is an early clinically informative diagnostic marker that often precedes clinical diagnosis (<u>Ahmed et al., 2013</u>; <u>Mueller et al.,</u> <u>2018</u>; <u>Slegers et al., 2018</u>). Across the AD continuum, language decline is driven by progressive lexical-semantic knowledge breakdown, reflected in gradually worsening naming, verbal fluency, and word knowledge (<u>García, 2026</u>; <u>Joubert et al., 2021</u>; <u>Macoir et al., 2019</u>). Standard neuropsychological batteries used in clinical practice and in large AD cohorts sample language mainly rely on verbal fluency and confrontation naming(<u>Mueller et al., 2005</u>; <u>National</u> <u>Alzheimer’s Coordinating Center, 2015</u>). Although informative, these tests have two important limitations. First, because they draw on both linguistic and non-linguistic processes, a low score cannot be unambiguously attributed to a specific language deficit (<u>Amunts et al., 2020</u>; <u>Henry et</u> <u>al., 2004</u>; <u>Kavé & Sapir-Yogev, 2020</u>). Second, they leave domains such as semantic memory, phonology and morphosyntax largely unexamined (<u>García, 2026</u>). As a result, it remains unclear which language components are affected, and at what stage, across the AD continuum.

Domain-specific language tests can address these limitations. The NACC frontotemporal lobar degeneration (FTLD) module is a battery developed to characterize the language features of individuals with frontotemporal lobar degeneration, and its subtests are organised by linguistic domain, each isolating a construct not sampled by the standard UDSv3 measures (<u>National</u> <u>Alzheimer’s Coordinating Center, 2015</u>). Although designed for FTLD phenotyping, this module presents an opportunity to compare *specialized* language tests across the AD continuum. Because the FTLD Module is administered within the same NACC protocol as the UDSv3, it uniquely enables a direct comparison of standard and specialized language tests within a single, biomarker-confirmed cohort. Unlike the standard tests, the FTLD Module tests are each designed to isolate a more specific linguistic process. They assess semantic memory (semantic association, word-picture matching), a domain not directly sampled by the UDSv3, as well as lexical retrieval (noun and verb naming), sentence-level phonological processing (sentence repetition and sentence reading), morphosyntax (Northwestern Anagram Test – Short Form), and single-word reading through both the lexical route (irregular words, which depend on stored whole-word representations) and the sublexical route (regular words, which can be read via grapheme-phoneme conversion), together with the standard measures forming a battery that samples language more comprehensively than either alone. Specialized tests therefore complement the standard tests for narrowing impairment to specific linguistic domains.

Nonetheless, few studies have comprehensively characterized language profiles across the AD spectrum using a broad range of language measures, including specialized tests such as those in the FTLD Module. Previous work has examined specific language domains or abilities in isolation: semantic memory (<u>Joubert et al., 2021</u>; <u>Montembeault et al., 2017</u>), lexical access to verbs (<u>Macoir et al., 2019</u>), morphosyntax (<u>Ahmed et al., 2013</u>; <u>Boschi et al., 2017</u>; <u>Ivanova et al., 2023a</u>; <u>Kavé & Goral, 2017</u>; <u>Mueller et al., 2018</u>; <u>Sung Jee et al., 2020</u>), phonological processing (<u>Kaltsa et al., 2024</u>), and single word reading (<u>Marier et al., 2024</u>), but a comprehensive assessment across domains in a large sample remains limited. Such an approach could inform diagnostic assessment by extending beyond commonly used measures such as verbal fluency and picture naming, while also identifying language domains that may be relevant for targeted intervention. Furthermore, with the recent expansion of the biological definition of AD, language profiles should be characterized in biomarker-confirmed samples. Most previous studies of MCI and AD have relied on clinical diagnoses without biological confirmation of AD pathology. Examining amyloid-biomarker-confirmed MCI and dementia due to AD may therefore provide a characterization more closely aligned with contemporary models of the disease.

To resolve these gaps, the present study aimed to evaluate the diagnostic utility of standard (UDSv3) and specialized (FTLD Module) language tests across the biologically confirmed AD clinical continuum (i.e., HC, MCI, and DAT). We hypothesized that specialized tests would contribute to the discrimination of AD stages, in addition to standard language tests. More specifically, semantic tests would provide the strongest discrimination at the earliest contrast (MCI vs. HC) consistent with the early and progressive vulnerability of semantic processing (<u>Joubert et al., 2021</u>). By contrast, we expected the other domains to show a more nuanced pattern. Phonological and morphosyntactic abilities are thought to be largely spared in MCI and the mild stage of DAT, yet deficits have been reported on more demanding tasks and tend to emerge with disease progression (<u>García, 2026</u>). Similarly, the contrast between relatively preserved rule-based and impaired memory-based processing in AD suggests that lexical route (irregular word reading) may be more vulnerable than sublexical route (regular word reading) (<u>García, 2026</u>). These domains may therefore show smaller or later-emerging differences, most evident in contrasts involving DAT. Because they have rarely been examined together within the same sample, their relative diagnostic value remains an open question.

## 2. Methods

This study is a cross-sectional secondary analysis of a cohort from the National Alzheimer’s Coordinating Center (NACC) database (www.naccdata.org). The NACC aggregates standardized clinical and neuropathological research data from NIA-funded Alzheimer’s Disease Research Centers (ADRCs) across the United States. While the NACC cohort is followed longitudinally, the present analyses were restricted to a single visit per participant to characterize language and clinical profiles at a specific stage of disease progression.

### 2.1 Participants

Participants were selected from the NACC, with visits conducted between March 2015 and March 2024 across 14 ADRCs. The final sample comprised 286 participants: 53 with MCI, 108 with DAT, and 125 cognitively unimpaired healthy control (HC) participants. Clinical diagnoses were established by ADRC consensus teams using relevant diagnostic criteria at the time of assessment (<u>Albert et al., 2011</u>; <u>McKhann et al., 2011</u>). The MCI group comprised amnestic and non-amnestic subtypes, both single- and multiple-domain, collapsed into the same participant group because of sample-size considerations. All DAT participants had a typical amnestic presentation.

To participate in the current study, we selected participants aged 45 years or older, who had English as their primary language, and who had completed all UDSv3 language tests and at least eight of nine FTLD Module language tests (<u>Weintraub et al., 2018</u>). This criterion was selected to balance data completeness with an adequate sample size, informed by sample size estimates under different missing-data thresholds. HC participants were required to meet criteria for normal cognition at the time of visit, operationalized as a global score of 0 on the CDR Dementia Staging Instrument plus NACC FTLD Behavior & Language Domains (hereafter CDR plus NACC FTLD) and race- and ethnicity-stratified MoCA cutoffs (i.e., ≤ 25 for non-Hispanic

White, ≤ 24 for Hispanic, ≤ 23 for non-Hispanic Black participants) (<u>Milani et al., 2018</u>; <u>Morris, 1993</u>; <u>Nasreddine et al., 2005</u>). DAT and MCI classification additionally required Core 1 biomarker positivity (amyloid-β, via positron emission tomography or cerebrospinal fluid) following the revised biomarker framework (<u>Jack et al., 2024</u>); HC participants were included without biomarker requirements. When multiple visits were available for a given participant, we retained a single visit, selected in order of priority: (1) availability of amyloid biomarker data, (2) number of completed tests. When multiple visits met the same criteria, we retained the most recent visit to ensure the accuracy of clinical diagnosis (<u>Koepsell & Monsell, 2012</u>; <u>Rasmusson et al., 1996</u>).

To address group size imbalances, HC participants were selected in two separate matching steps: controls were matched to the MCI group and, independently, to the DAT group, on age and education (nearest available), with exact matching on sex. The two matched HC sets were then merged, with duplicates removed, to form the single HC group used in all analyses.

All patients were retained; only HC were downsampled, yielding 108 DAT, 53 MCI, and 125 HC. All participants or their informants provided written informed consent at their respective ADRCs. Institutional Review Board approvals were obtained by the NACC from all participating institutions.

### 2.2 Procedure

#### 2.2.1 Language Assessment

We included the three standard language tasks from the NACC UDSv3: semantic fluency (animals and vegetables), letter fluency (F and L), and confrontation naming from the Multilingual Naming Test (MINT). To further characterize specific linguistic profiles, nine specialized tasks from the FTLD Module were also analyzed: regular and irregular word reading, semantic word-picture matching, semantic associates, sentence construction task from the Northwestern Anagram Test (NAT-S), noun and verb naming from the Northwestern Naming Battery (NNB), sentence repetition, and sentence reading (<u>Gefen et al., 2020</u>; <u>Gollan et al., 2012</u>; <u>Thompson et al., 2012</u>; <u>Weintraub et al., 2018</u>; <u>Weintraub et al., 2009</u>). An overview of the language assessment battery is provided in Table 1.

**Table 1.** Overview of Standard (UDSv3) and Specialized (FTLD Module) Language Tasks.

| Test | Test description | Primary language domain assessed |
| --- | --- | --- |
| *Standard* | *UDSv3 core language measures* |  |
| Semantic verbal fluency | Generate as many category members (animals or vegetables) as possible in 60 seconds | **Semantic memory organization** and associative lexical retrieval |
| Phonemic verbal fluency | Generate as many words as possible beginning with a specified letter (“F” or “L”) in 60 seconds | **Controlled lexical retrieval, inhibition** of semantic associates, **set maintenance** and **switching** |
| Confrontation naming (MINT) | Name pictured objects (32 items) | **Semantic-lexical retrieval** (oral production) |
| *Specialized* | *FTLD Module addition* |  |
| Noun naming (NNB subtest) | Name pictured objects (16 noun items) as quickly as possible | **Semantic-lexical retrieval** (oral production) |
| Verb naming (NNB subtest) | Name pictured actions (16 verb items) as quickly as possible | **Semantic-lexical retrieval** (oral production) with heavier load on event/action semantics and morphosyntactic features (verb argument structure) compared to noun naming |
| Word-picture matching | Point to the picture corresponding to a spoken word among four semantically related (20 trials). | **Lexical-semantic access** and semantic discrimination/executive selection among foils (oral comprehension) |
| Semantic associates | Point to the picture pair that is semantically related from two simultaneously presented picture pairs (16 trials). | **Semantic memory**, especially conceptual relations/associations (amodal semantic hub) |
| Sentence repetition | Repeat sentences of varying in length and syntactic complexity (5 items) | **Phonological buffer** for multiword sequences |
| Sentence reading | Read aloud the same 5 sentences as in the repetition task (5 items) | **Phonology**; grapheme to phoneme conversion |
| Sentence construction (NAT-S) | Construct grammatically correct sentences from printed word cards to match a picture (10 items: 5 canonical subject who-questions and 5 non-canonical object who-questions) | **Morphosyntax**; ordering constituents and thematic role assignment |
| Regular word reading | Read aloud words with regular grapheme–phoneme correspondences (15 items). | **Sublexical** (grapheme to phoneme conversion) reading route |
| Irregular word reading | Read aloud irregularly spelled words that violate standard grapheme–phoneme correspondences (15 items). | **Lexical-semantic** reading route |

*Note.* UDSv3 = Uniform Dataset version 3 (Weintraub et al., 2018). The semantic (automatic) versus phonemic (controlled) retrieval characterization of the verbal fluency tasks follows Marko et al. \(2023\). MINT = Multilingual Naming Test (Ivanova et al., 2013). FTLD module = Frontotemporal Lobar Degeneration Module (National Alzheimer’s Coordinating Center, 2015). NNB = Northwestern Naming Battery (Thompson et al., 2012). NAT-S = Northwestern Anagram Test — Short Form (Weintraub et al., 2009). Two derived measures are computed from the tasks above.

#### 2.2.2 Cognitive Functions

Dementia severity was staged with the CDR plus NACC FTLD, whose global score is 0 (no impairment), 0.5 (very mild/questionable), 1 (mild), 2 (moderate), or 3 (severe) (Morris, 1993).

Global cognition was screened with the Montreal Cognitive Assessment (MoCA), a brief 30-point measure on which higher scores indicate better performance (<u>Nasreddine et al., 2005</u>).

### 2.3 Statistical analyses

To assess group differences across language tests, we conducted one-way analyses of covariance (ANCOVAs) comparing HC, MCI, and DAT groups on each language measure, with age, sex, and education as covariates; pairwise comparisons used Tukey’s HSD to correct for multiple comparisons. To evaluate the diagnostic performance of individual language tests, we computed receiver operating characteristic (ROC) curves and AUC for each test across three pairwise classification problems (i.e., DAT vs. HC, MCI vs. HC, and DAT vs. MCI). We defined clinically meaningful diagnostic accuracy as AUC ≥ .700 (<u>Corbacioglu & Aksel, 2023</u>; <u>Huque et</u> <u>al., 2025</u>). Optimal cutoffs were determined using Youden’s index (<u>Youden, 1950</u>). AUC, sensitivity, and specificity at the optimal cutoff are reported with 95% confidence intervals derived from 2,000 stratified bootstrap resamples. Lastly, to compare diagnostic performance between standard UDSv3 and specialized FTLD Module tests, differences in AUC were evaluated using DeLong’s test (<u>DeLong et al., 1988</u>). All statistical analyses were conducted using R 4.4.3 (<u>R Core Team, 2025</u>).

## 3. Results

### 3.1 Demographics

Demographic and clinical characteristics are presented in Table 2. Age differed across groups, *F*(2, 283) = 10.94, *p* < .001, *η²* = .072. The MCI group was older than both HC (*p* < .001) and DAT (*p* < .001), whereas HC and DAT did not differ (*p* = .92). Age was therefore included as a covariate in all analyses. No significant differences were found for education, *F*(2, 283) = 0.82, *p* = .442, *η²* = .006, or sex, *χ²*(2) = 0.48, *p* = .786. MoCA scores declined systematically across groups, *F*(2, 276) = 356.28, *p* < .001, *η²* = .721. Both CDR, *F*(2, 283) = 365.50, *p* < .001, *η²* = .721, and disease duration, *F*(2, 283) = 159.02, *p* < .001, *η²* = .529, increased. Dementia severity in the DAT group was predominantly mild (mean CDR = 0.9 ± 0.4) and questionable for MCI (mean CDR = 0.5 ± 0.1) (<u>Morris, 1993</u>).

**Table 2.** Demographic and Clinical Characteristics of Participants.

| **Variable** | **HC** | **MCI** | **DAT** |
| --- | --- | --- | --- |
| Sample size (n) | 125 | 53 | 108 |
| ***Demographic characteristics*** |  |  |  |
| Mean age (years) | 60.2 ±8.0 | 65.0 ±7.8 | 59.9 ±4.7 |
| Age range (years) | 47 - 86 | 48 - 84 | 49 - 75 |
| Sex (%) | M = 46.4, F = 53.6 | M = 49.1, F = 50.9 | M = 50.9, F = 49.1 |
| Years of education | 15.9 ±2.1 | 16.2 ±2.4 | 15.7 ±2.7 |
| ***Clinical characteristics*** |  |  |  |
| Global CDR | 0.0 ±0.0 | 0.5 ±0.1 | 0.9 ±0.4 |
| Disease duration (years) | 0.0 ±0.0 | 4.7 ±3.0 | 4.9 ±3.1 |
| Global cognition - MoCA (/30) | 28.0 ±1.5 | 19.9 ±5.3 | 13.2 ±5.6 |
| ***Ethnicity (%)*** |  |  |  |
| White | 92.0 | 96.2 | 91.7 |
| Black or African American | 4.8 | 3.8 | 6.5 |
| American Indian or Alaska Native | 0.0 | 0.0 | 0.0 |
| Native Hawaiian or Other Pacific Islander | 0.0 | 0.0 | 0.9 |
| Asian | 2.4 | 0.0 | 0.9 |
| Other | 0.0 | 0.0 | 0.0 |
| Unknown | 0.8 | 0.0 | 0.0 |
| ***Biomarkers status*** |  |  |  |
| *Amyloid (%)* |  |  |  |
| Amyloid + | 0.0 | 100 | 100 |
| Amyloid - | 7.2 | 0.0 | 0.0 |
| n/a | 92.8 | 0.0 | 0.0 |
| *Tau in PET scan (%)* |  |  |  |
| Tau + | 0.0 | 28.3 | 49.1 |
| Tau - | 4.8 | 9.4 | 0.9 |
| n/a | 95.2 | 62.3 | 50.0 |
| *Tau in cerebrospinal fluid (%)* |  |  |  |
| Tau + | 0.0 | 30.2 | 28.7 |
| Tau - | 0.8 | 9.4 | 4.6 |
| n/a | 99.2 | 60.4 | 66.7 |

*Note.* HC = healthy controls; MCI = mild cognitive impairment; DAT = Dementia of Alzheimer's type. M= male. F = Female. n/a = Unavailable biomarker. Global CDR = global score on the CDR Dementia Staging Instrument plus NACC FTLD Behavior & Language Domains. MoCA = Montreal Cognitive Assessment (Nasreddine et al., 2005).

### 3.2 Descriptive Statistics of Language Tests across Groups

Descriptive statistics and between-group comparisons for both standard and specialized language measures are presented in Table 3. Groups showed significant differences across the standard language tests. Semantic fluency demonstrated the largest effect, *F*(2, 280) = 236.37, *p* < .001, *η²* = .628, followed by letter fluency, F(2, 280) = 55.05, *p* < .001, *η²* = .282, and MINT, *F*(2, 280) = 39.86, *p* < .001, *η²* = .222. Among standard tests, semantic and letter fluency followed a progressive decline pattern (DAT < MCI < HC), whereas MINT differentiated both patient groups from HC but not from each other (DAT = MCI < HC).

**Table 3.**
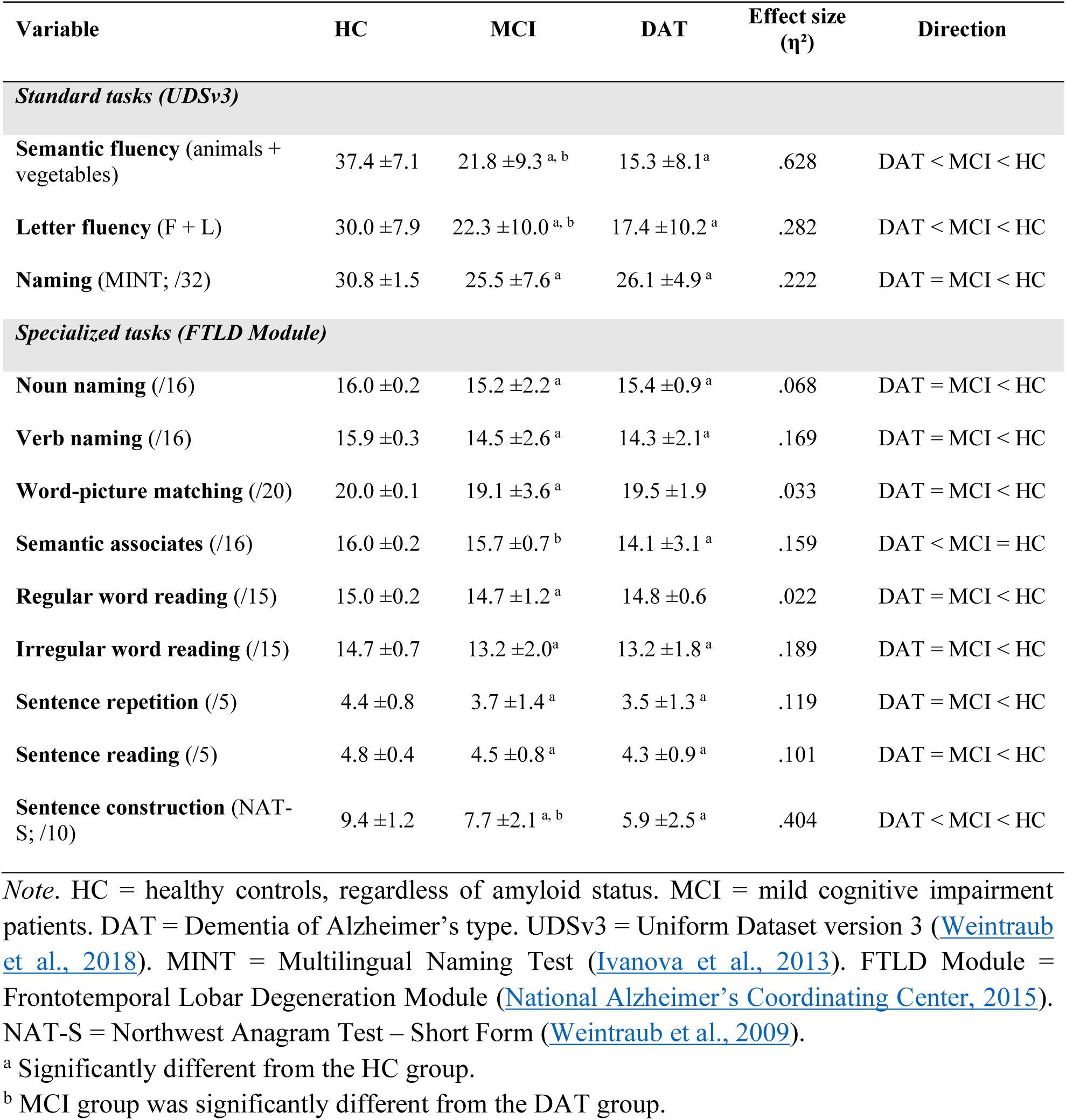
Descriptive Statistics and Post-Hoc Test Results for Language Performance.

| Variable | HC | MCI | DAT | Effect size ($\eta^2$) | Direction |
| --- | --- | --- | --- | --- | --- |
| *Standard tasks (UDSv3)* |  |  |  |  |  |
| **Semantic fluency** (animals + vegetables) | 37.4 ± 7.1 | 21.8 ± 9.3 <sup>a, b</sup> | 15.3 ± 8.1<sup>a</sup> | .628 | DAT < MCI < HC |
| **Letter fluency** (F + L) | 30.0 ± 7.9 | 22.3 ± 10.0 <sup>a, b</sup> | 17.4 ± 10.2 <sup>a</sup> | .282 | DAT < MCI < HC |
| **Naming** (MINT; /32) | 30.8 ± 1.5 | 25.5 ± 7.6 <sup>a</sup> | 26.1 ± 4.9 <sup>a</sup> | .222 | DAT = MCI < HC |
| *Specialized tasks (FTLD Module)* |  |  |  |  |  |
| **Noun naming** (/16) | 16.0 ± 0.2 | 15.2 ± 2.2 <sup>a</sup> | 15.4 ± 0.9 <sup>a</sup> | .068 | DAT = MCI < HC |
| **Verb naming** (/16) | 15.9 ± 0.3 | 14.5 ± 2.6 <sup>a</sup> | 14.3 ± 2.1<sup>a</sup> | .169 | DAT = MCI < HC |
| **Word-picture matching** (/20) | 20.0 ± 0.1 | 19.1 ± 3.6 <sup>a</sup> | 19.5 ± 1.9 | .033 | DAT = MCI < HC |
| **Semantic associates** (/16) | 16.0 ± 0.2 | 15.7 ± 0.7<sup>b</sup> | 14.1 ± 3.1 <sup>a</sup> | .159 | DAT < MCI = HC |
| **Regular word reading** (/15) | 15.0 ± 0.2 | 14.7 ± 1.2 <sup>a</sup> | 14.8 ± 0.6 | .022 | DAT = MCI < HC |
| **Irregular word reading** (/15) | 14.7 ± 0.7 | 13.2 ± 2.0<sup>a</sup> | 13.2 ± 1.8 <sup>a</sup> | .189 | DAT = MCI < HC |
| **Sentence repetition** (/5) | 4.4 ± 0.8 | 3.7 ± 1.4 <sup>a</sup> | 3.5 ± 1.3 <sup>a</sup> | .119 | DAT = MCI < HC |
| **Sentence reading** (/5) | 4.8 ± 0.4 | 4.5 ± 0.8 <sup>a</sup> | 4.3 ± 0.9 <sup>a</sup> | .101 | DAT = MCI < HC |
| **Sentence construction** (NAT-S; /10) | 9.4 ± 1.2 | 7.7 ± 2.1 <sup>a, b</sup> | 5.9 ± 2.5 <sup>a</sup> | .404 | DAT < MCI < HC |

*Note.* HC = healthy controls, regardless of amyloid status. MCI = mild cognitive impairment patients. DAT = Dementia of Alzheimer's type. UDSv3 = Uniform Dataset version 3 (Weintraub et al., 2018). MINT = Multilingual Naming Test (Ivanova et al., 2013). FTLN Module = Frontotemporal Lobar Degeneration Module (National Alzheimer's Coordinating Center, 2015). NAT-S = Northwest Anagram Test – Short Form (Weintraub et al., 2009).
<sup>a</sup> Significantly different from the HC group.
<sup>b</sup> MCI group was significantly different from the DAT group.

Specialized FTLD Module tests showed more variable group patterns. A progressive decline pattern (DAT < MCI < HC) was observed for the NAT-S total score, *F*(2, 263) = 89.04, *p* < .001, *η²* = .404. The semantic associates test showed DAT < MCI = HC (*η²* = .159).

Sentence-level phonological tests showed DAT and MCI both differing from HC but not from each other: sentence repetition, *F*(2, 279) = 18.77, *p* < .001, *η²* = .119, and sentence reading, *F*(2, 280) = 15.72, *p* < .001, *η²* = .101, both differentiated DAT and MCI from HC (*p* ≤ .006) but not from each other (*p* = .524 and *p* = .485, respectively). Irregular word reading, noun naming, and verb naming showed the same pattern (DAT = MCI < HC; *η²* = .068–.189). For regular word reading and word-picture matching, only the HC–MCI difference was significant (p = .047 and p = .012, respectively); DAT differed from neither group. Both effects were negligible in magnitude (η² = .022 and .033). Figure 1 shows the score distributions and modal scores for semantic fluency (Panel A1) and the NAT-S total score (Panel B1). The corresponding plots for sentence reading and sentence repetition appear in Supplementary Figure S1.

**Figure 1.**
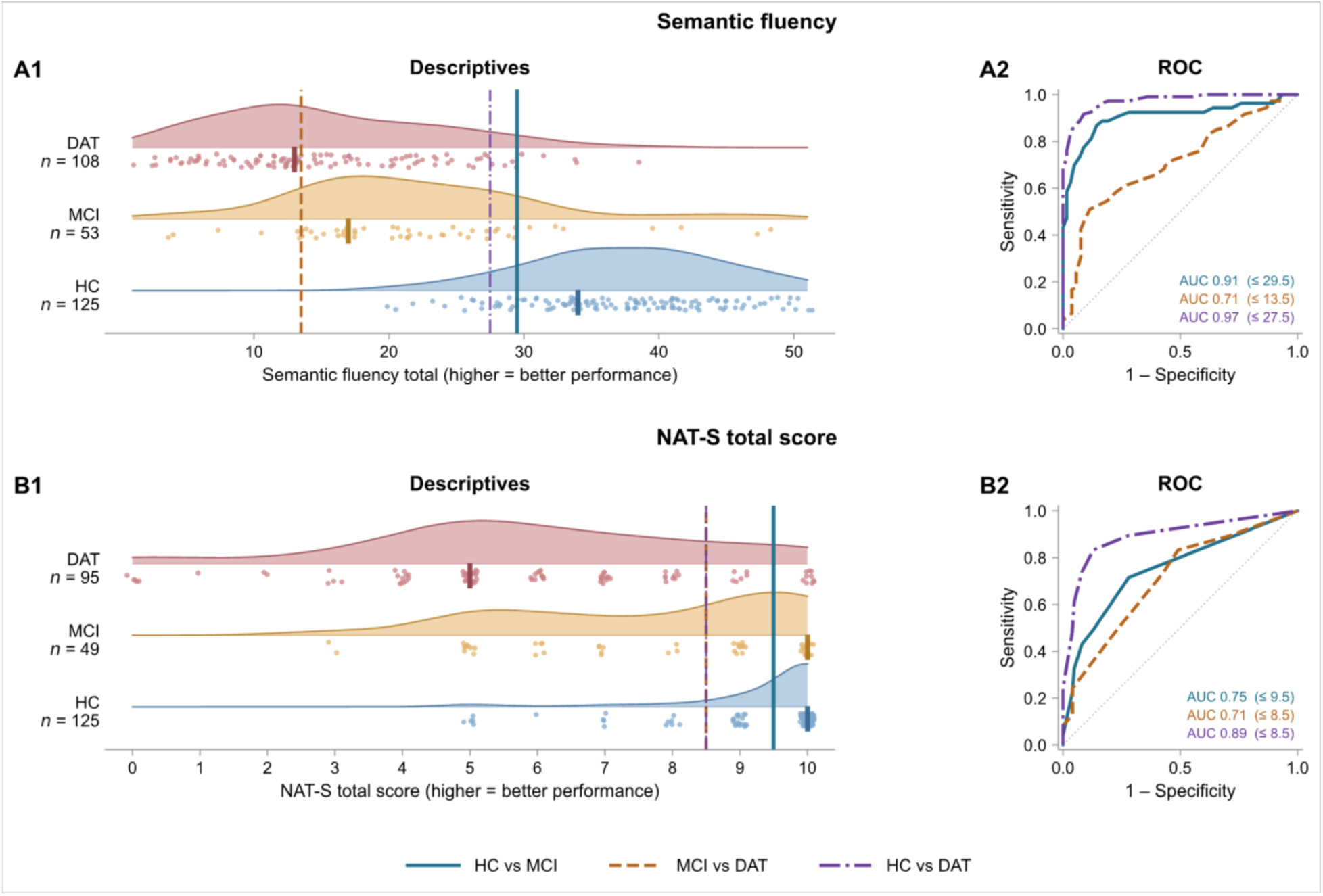
Score Distributions and ROC Curves for Semantic Fluency and NAT-S Total Score. *Note*. **A1 and A2** show semantic fluency; **B1 and B2** show NAT-S total score. **Panels A1–B1**: shaded curves are kernel density estimates, points are individual scores (jittered), and the vertical bar marks each group’s modal score. **Vertical reference lines** mark the Youden J-optimal cut-off for each pairwise comparison, matched by colour and line style to the curves in Panels A2–B2. **Panels A2–B2**: the diagonal indicates chance performance. AUC and the corresponding Youden cut-off are printed within each ROC panel, colour-matched to the contrast. For NAT-S total score, the MCI–DAT and HC–DAT cut-offs **coincide** at 8.5. AUC = area under the curve, with 95% bootstrap confidence intervals (2,000 resamples). NAT-S total score = Northwestern Anagram Test – Short Form. HC = healthy control; MCI = mild cognitive impairment; DAT = Alzheimer’s disease dementia.

### 3.3 Diagnostic Performance of Standard and Specialized Measures

AUC values, optimal cutoffs, sensitivities, and specificities for all language measures across three diagnostic comparisons are presented in Supplementary Table 1. Figure 1 shows the pairwise diagnostic performance of semantic fluency, NAT-S total score, sentence reading, and sentence repetition across the groups. DeLong test results are presented in Supplementary Table 2.

#### 3.3.1 DAT vs. HC

Eight out of twelve tests demonstrated clinically meaningful diagnostic accuracy (i.e., AUC ≥ .700). Semantic fluency showed the highest discrimination, followed by NAT-S total score, MINT, letter fluency, verb naming, irregular word reading, sentence repetition, and semantic associates. All other measures fell below the AUC threshold. Semantic fluency significantly outperformed letter fluency, MINT, and all specialized tests (ΔAUC = .08–.43, *z* = 3.58–25.12, all *p* ≤ .017). NAT-S total score was the strongest specialized measure, exceeding every other test (ΔAUC = .11–.34, *z* = 3.40–13.20, all *p* ≤ .033) except letter fluency (ΔAUC = .06, *z* = 1.91, *p* = 1.00) and MINT (ΔAUC = .03, *z* = 0.84, *p* = 1.00), from which it did not differ; semantic fluency in turn exceeded NAT-S (ΔAUC = .08, *z* = 3.58, *p* = .017).

#### 3.3.2 MCI vs. HC

Five out of twelve language tests achieved clinically meaningful diagnostic accuracy. Semantic fluency again showed the strongest performance, followed by MINT, NAT-S total score, letter fluency, and irregular word reading. Semantic fluency significantly outperformed nine of the eleven remaining measures (ΔAUC = .16–.37, *z* = 4.10–10.64, all *p* ≤ .003) but did not differ from MINT (ΔAUC = .13, *z* = 3.17, *p* = .112) or NAT-S total score (ΔAUC = .15, *z* = 3.14, *p* = .119). The remaining clinically meaningful measures did not differ from one another (all *p* = 1.00).

#### 3.3.3 DAT vs. MCI

Only two tests achieved clinically meaningful diagnostic accuracy for this challenging boundary: semantic fluency and NAT-S total score. All other measures fell below the .700 threshold. Semantic fluency significantly exceeded regular word reading, irregular word reading, noun naming, word-picture matching, MINT, and sentence reading (ΔAUC = .17–.21, *z* = 3.69–4.87, all *p* ≤ .022), but did not differ from the remaining measures.

### 3.4 Post-Hoc Analysis: Executive Functions Partly Account for Morphosyntactic Performance

Among the specialized language tests, the NAT-S was the only measure to show acceptable discrimination across all three contrasts, and, along with semantic fluency, the only one to do so for DAT vs. MCI, with performance declining progressively across groups (DAT < MCI < HC). This graded decline was unexpected: we had hypothesized that morphosyntactic measures would be insensitive to the MCI vs. HC distinction, given the view that morphosyntax is relatively preserved early in the disease (<u>Kavé & Goral, 2017</u>). Morphosyntactic processing and executive functions are, however, closely linked. Arranging printed word cards into a grammatical sentence requires holding the constituents in mind and reordering them, so the task draws on both (<u>Fyndanis et al., 2022</u>; <u>Ivanova et al., 2023a</u>). A low score on this task could therefore reflect a morphosyntactic deficit, an executive deficit, or both, since executive deficits were well documented in DAT (<u>Guarino et al., 2018</u>).

To address this, we used the two executive measures available in the UDSv3. In the Trail Making Test Part B (TMT-B; <u>Reitan, 1958</u>; <u>Weintraub et al., 2018</u>), participants alternate between numbers and letters in sequential order as quickly as possible, with completion time indexing cognitive flexibility (longer times indicate poorer performance). In the Number Span Test: Backward, participants repeat digit sequences in reverse order, with the number of correct trials (0–14) indexing working memory (<u>Weintraub et al., 2018</u>).

To test whether group differences on the NAT-S persisted after accounting for executive function and working memory, we fitted a hierarchical linear regression on NAT-S score: Step 1 entered age, sex, education, and diagnosis (dummy-coded, HC as reference), and Step 2 added TMT-B and the Number Span Test: Backward.

The model was fitted with 216 participants (125 HC, 44 MCI, 47 DAT) due to missing TMT-B data (57 DAT, 7 MCI). Excluded DAT participants were more cognitively impaired than those retained (MoCA: 10.2 vs. 17.0, *d* = 1.56), indicating non-random attrition with respect to disease severity. Multicollinearity was acceptable (adjusted generalized variance inflation factors; range: 1.01-1.70; Supplementary Table 3) (<u>Fox & Monette, 1992</u>; <u>Kim, 2019</u>).

In Step 1, NAT-S score differed by group (MCI: *B* = -1.63, *p* < .001; DAT: *B* = -2.19, *p* < .001; *R²* = .261). Adding cognitive flexibility and working memory increased explained variance, *ΔR²* = .096, F(2, 208) = 15.56, *p* < .001. Longer TMT-B completion time (*B* = -.008, *p* < .001) and lower backward span (*B* = .12, *p* = .036) were each associated with lower NAT-S scores. From Step 1 to Step 2, the MCI coefficient changed from *B* = -1.63 to *B* = -.73 (55.5% reduction) and the DAT coefficient from *B* = -2.19 to *B* = -.58 (73.4% reduction); group differences were attenuated, and the DAT difference was no longer significant while the MCI difference remained significant (MCI: *p* = .028; DAT: *p* = .157). Model summaries are presented in Supplementary Tables 4 and 5. Figure 2 shows the standardized coefficients.

**Figure 2.**
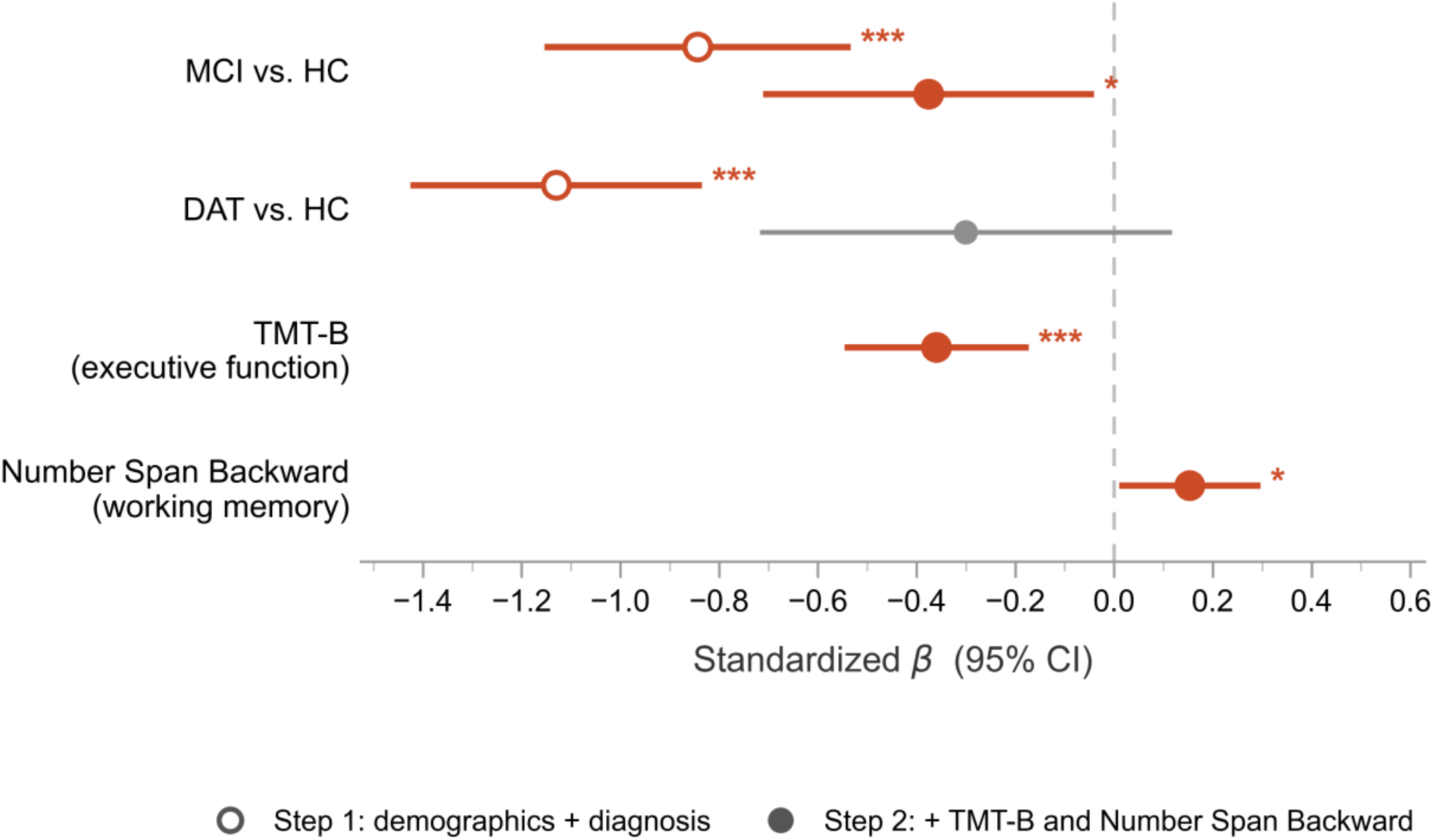
Standardized Coefficients from the Hierarchical Regression Predicting NAT-S Total Score. *Note.* Points are standardized *β* with 95% CIs. Open circles show Step 1 (age, sex, education, and diagnosis); filled circles show Step 2, which added TMT-B and the Number Span Test: Backward. TMT-B = Trail Making Test Part B (executive function). * = *p* < .05, ** = *p* < .01, *** = *p* < .001.

## 4. Discussion

### 4.1 Summary of findings

In this study, we aimed to establish comprehensive language profiles across the biomarker-confirmed AD continuum and to evaluate the diagnostic utility of standard (UDSv3) and specialized (FTLD Module) language tests for distinguishing MCI and DAT from HC and from each other. Our hypothesis that specialized tests would contribute to discrimination beyond standard tests was partly supported. Semantic fluency, a standard test, and the NAT-S, a specialized morphosyntactic test, were the only tests to reach clinically meaningful accuracy in all three contrasts. The performance of the NAT-S was unexpected, as we had anticipated morphosyntactic differences to emerge mainly at the dementia stage. The strong performance of semantic fluency and NAT-S may partly reflect their reliance on executive functioning (<u>Amunts</u> <u>et al., 2020</u>; <u>Henry et al., 2004</u>), as our post hoc analysis supports for the NAT-S. MINT and letter fluency also reached clinically meaningful accuracy in distinguishing MCI and DAT from HC, but not in distinguishing DAT from MCI. Irregular word reading was the only other specialized measure to reach clinically meaningful accuracy in distinguishing both MCI and DAT from HC. The remaining specialized tests of semantic processing, lexical retrieval, phonology, and sublexical reading reached clinically meaningful accuracy only for DAT vs. HC, or not at all. Overall, these results support the clinical utility of standard language tests while highlighting the potential diagnostic value of selected specialized tests.

### 4.2. Most Accurate Language Tests Across the AD Continuum: Semantic Verbal Fluency and NAT-S

Semantic verbal fluency showed the highest overall accuracy across all three diagnostic contrasts (HC vs. MCI vs. DAT), in line with previous literature (<u>Henry et al., 2004</u>; <u>McDonnell et al., 2020</u>). This supports its widespread use in clinical and research protocols in AD (<u>National Alzheimer’s Coordinating Center, 2015</u>; <u>Petersen et al., 2010</u>), despite an important limitation: performance on this task depends on both linguistic and non-linguistic processes, such that a low score cannot be unambiguously attributed to a specific language deficit (<u>Amunts et al., 2020</u>; <u>Henry et al., 2004</u>; <u>Kavé & Sapir-Yogev, 2020</u>). Indeed, verbal fluency requires participants to generate and retrieve concepts, inhibit responses from competing semantic categories, monitor previously produced words, and strategically organize their search (e.g., generating marine animals followed by birds). Nevertheless, our findings suggest that semantic verbal fluency remains highly clinically informative despite its multidimensional nature. It draws on both semantic memory and executive functions, which are affected in a graded manner along the AD continuum and may contribute to the prominent diagnostic performance (<u>Joubert et al., 2021</u>; <u>McDonnell et al., 2020</u>). More detailed approaches have been proposed to isolate the semantic processes underlying performance on this task, including item-level analyses of the semantic complexity of generated words and analyses of clustering (<u>De Marco et al., 2025b</u>), whereby larger cluster sizes may reflect more efficient retrieval of exemplars within semantic subcategories (<u>De Marco et al., 2025a</u>). Such approaches could provide greater insight into the specific semantic processes contributing to verbal fluency performance while preserving the clinical practicality of this widely used test.

The NAT-S was the only specialized test to reach clinically meaningful accuracy in all three contrasts. Contrary to the view that syntax is largely spared in early AD, this aligns with growing connected speech evidence that syntactic simplification may emerge as early as the MCI stage (<u>Ivanova et al., 2023a</u>; <u>Sung Jee et al., 2020</u>). Two competing accounts have been proposed regarding the origin of such morphosyntactic deficits in dementia. The lexical-semantic origin hypothesis attributes it to degraded lexical-semantic representations, whereas the cognitive origin hypothesis attributes it to the decline of domain-general cognitive functions such as executive function and working memory (<u>Ivanova et al., 2023a</u>). Because the NAT-S supplies the lexical items and is untimed, it minimizes lexical-retrieval, speed, and articulatory demands, allowing the two hypotheses to be tested after adjusting for TMT-B and backward digit span (<u>Boschi et al., 2017</u>; <u>Ivanova et al., 2023b</u>; <u>Mueller et al., 2018</u>; <u>Weintraub et al., 2009</u>). The post-hoc analysis suggests that executive functions account for a large part of the morphosyntactic decline. TMT-B and backward span each independently predicted NAT-S performance, and once they were entered the group coefficients were reduced by 55.5% (MCI) and 73.4% (DAT). The DAT difference was no longer significant, consistent with the cognitive origin hypothesis, whereas the MCI difference remained significant, which may reflect a morphosyntactic component partly independent of domain-general resources.

Two limitations should be considered when interpreting these findings. First, the NAT-S samples a narrow range of syntactic structures and does not assess others such as passives and relative clauses (<u>Bickel et al., 2000</u>). Consequently, findings cannot be generalized to syntax in unconstrained speech. Second, the DAT sample was reduced by 56.5% (from 108 to 47), primarily because of missing TMT-B scores. Because TMT is less likely to be completed at greater disease severity, the retained participants were likely those with milder impairment, which may have attenuated the group difference.

### 4.3. Language Tests with Greater Accuracy for Early HC–MCI Distinction: Letter Fluency, MINT, and Irregular Word Reading

Letter fluency and MINT reached clinically meaningful accuracy in distinguishing MCI and DAT from HC, but not DAT from MCI, suggesting that they are sensitive to early impairment but of limited use for staging. MINT performance was similar in MCI and DAT, whereas letter fluency differed between them at the group level but with too much overlap to reach clinically meaningful accuracy.

As hypothesized, single-word reading was impaired through the lexical-semantic route only: irregular word reading reached clinically meaningful accuracy for MCI vs. HC and DAT vs. HC, whereas regular word reading, which relies on the sublexical route, did not reach it in any contrast (García, 2026). Because irregular words cannot be read by grapheme–phoneme conversion, they depend on stored lexical and semantic knowledge, which is vulnerable early in AD (Joubert et al., 2021). With the NAT-S, irregular word reading was thus the only specialized test to distinguish MCI from HC, suggesting that it may capture early lexical-semantic vulnerability.

The remaining specialized tests were informative mainly at the dementia stage. Phonological tests reached clinically meaningful accuracy only for DAT vs. HC (sentence repetition) or not at all (sentence reading), consistent with previous literature describing phonology as a domain relatively spared until later stages (<u>García, 2026</u>). Among lexical retrieval tests, verb naming reached clinically meaningful accuracy for DAT vs. HC whereas noun naming did not, consistent with a greater deficit of verb naming in DAT (<u>Macoir et al., 2019</u>). Among semantic memory tests, semantic associates reached this threshold for DAT vs. HC only, and word-picture matching in no contrast; contrary to our hypothesis, neither discriminated MCI from HC.

Apart from semantic fluency, standard measures remain sensitive in detecting early impairment but did not differentiate DAT from MCI, whereas most specialized measures were informative mainly at the dementia stage by detecting specific impairments.

### 4.4. Specialized Language Tests: Potential Clinical Utility Despite Limited Group-Level Accuracy

These results do not imply that specialized measures were not adequate for clinical use. First, the AUC threshold alone may not capture their utility. HC performed at ceiling on regular word reading, word-picture matching, semantic associates, and noun naming, such that the optimal cutoff in contrasts against HC classified a single error as impaired, yielding high specificity (.96-.99) but low sensitivity (.09-.44). These specialized tests thus produced few false positives, but they also often missed impaired participants. This pattern may explain why semantic memory tests did not discriminate MCI from HC as hypothesized. The diagnostic utility of these language domains therefore cannot be fully determined from the present data.

Second, similar AUCs can conceal different error profiles. In distinguishing DAT from MCI, the NAT-S and semantic fluency reached similar accuracy (AUC = .708 and .710 respectively) but showed opposite profiles of sensitivity and specificity at their optimal cutoffs. NAT-S was sensitive (.832) but not specific (.510), whereas semantic fluency was specific (.887) but not sensitive (.509). This suggests that the two tests may be complementary for staging along the AD continuum, although their combined accuracy was not tested in the present study.

Finally, while potentially less sensitive at the group level for detecting AD-related changes or differentiating disease stages across the broader AD spectrum, it is important to recognize that language deficits are highly heterogeneous in DAT and MCI: MCI subtypes differ in their language profiles (<u>Liampas et al., 2022</u>), and performance across standard language tests varies between early- and late-onset DAT independently of dementia severity (<u>Gallée et al.,</u> <u>2025</u>). These specialized language tests therefore remain clinically valuable for characterizing individual patients’ specific language profiles and identifying case-specific impairments that may not be captured by broader measures.

### 4.5 Limitations

Beyond the measure-specific limitations discussed above, several general limitations related to the study population apply to all analyses. First, the study was cross-sectional and cannot infer about progression, and optimal cutoffs and AUCs were derived and evaluated in the same sample without cross-validation. Accuracy estimates may be optimistic and should be replicated in an independent cohort. Second, because HC participants were included without biomarker requirements due to data unavailability, the HC group may include individuals in the preclinical stage of AD (i.e. amyloid-positive), which would likely attenuate group differences and render the present diagnostic accuracy estimates conservative. Future studies could compare amyloid-negative and amyloid-positive HC to identify preclinical changes. Second, the composition of the NACC sample limits the generalizability of these findings to broader populations. Participants were predominantly White (91.7–96.2% across groups) and highly educated (mean 15.7–16.2 years of education across groups), and therefore may not be representative of the broader, more diverse population of older adults. Third, MCI and DAT are clinically heterogeneous in their language deficits, and MCI subtypes were pooled in the present study to preserve sample size. In another cohort of 1,924 older adults, amnestic MCI performed worse than non-amnestic MCI, especially on semantic fluency and verbal comprehension, and this difference was driven by the multi-domain amnestic subgroup (<u>Liampas et al., 2022</u>).

Pooling subtypes may therefore have obscured subtype-specific language profiles in the MCI group. Finally, the findings should be interpreted in the context of the atypical age profile of the clinical samples. A notable characteristic of the DAT group is its relatively young age (M = 59.9 ± 4.7, range = 49–75;), such that the MCI group (M = 65.0 ± 7.8, range = 48–84) was, on average, older than the DAT group, a pattern opposite to the age gradient typically expected along the AD continuum. Although age was included as a covariate in ANCOVAs and hierarchical regression, ROC analyses were not age-adjusted and residual age-related confounding cannot be fully excluded, particularly in the DAT vs. MCI contrast. The relatively young age of the DAT group likely reflects the selective administration of the optional FTLD Module within NACC, which may be preferentially administered to younger patients. Because DAT participants were amyloid-positive, presented with a typical amnestic profile, and were age-matched to HC, this is unlikely to affect diagnostic accuracy relative to HC. Nevertheless, early-onset AD may differ from late-onset AD in its cognitive and linguistic profile (<u>Gallée et</u> <u>al., 2025</u>), and generalizability to typical late-onset AD populations should be confirmed in future work.

### 4.6 Conclusion

In a biomarker-confirmed MCI and DAT cohort, semantic fluency, a standard UDSv3 test, showed the highest accuracy across the continuum, although it did not significantly outperform NAT-S for MCI vs. HC or DAT vs. MCI. Among the specialized FTLD Module tests, morphosyntax (NAT-S) was the most informative domain, challenging the view that syntax is spared in early AD. Executive function partly explained morphosyntactic performance, suggesting a mainly domain-general contribution in DAT and a residual morphosyntactic component in MCI. Irregular word reading also distinguished MCI from HC, whereas the other standard and specialized tests detected impairment relative to HC without distinguishing DAT from MCI. Several specialized tests were limited by ceiling effects. Clinically, the NAT-S may complement semantic fluency for staging, and specialized tests may help characterize individual language profiles. Longitudinal studies with a broader range of syntactic and executive measures are needed to confirm these contributions and the incremental diagnostic value of the NAT-S.

## Data Availability

All data produced in the present study are available upon reasonable request to the authors

## Acknowledgement

The NACC database is funded by NIA/NIH Grant U24 AG072122. NACC data are contributed by the NIA-funded ADRCs: P30 AG062429 (PI James Brewer, MD, PhD), P30 AG066468 (PI Oscar Lopez, MD), P30 AG062421 (PI Teresa Gomez-Isla, MD), P30 AG066509 (PI Thomas Grabowski, MD), P30 AG066514 (PI Mary Sano, PhD), P30 AG066530 (PI Helena Chui, MD, Arthur Toga, PhD), P30 AG066507 (PI Marilyn Albert, PhD), P30 AG066444 (PI David Holtzman, MD), P30 AG066518 (PIs Lisa Silbert, MD, Kevin Duff, PhD), P30 AG066512 (PI Thomas Wisniewski, MD), P30 AG066462 (PI Scott Small, MD), P30 AG072979 (PI David Wolk, MD), P30 AG072972 (PIs Charles DeCarli, MD, Rachel Whitmer, PhD), P30 AG072976 (PI Andrew Saykin, PsyD), P30 AG072975 (PI Julie Schneider, MD, MS), P30 AG072978 (PI Ann McKee, MD), P30 AG072977 (PI Robert Vassar, PhD), P30 AG066519 (PI Joshua Grill, PhD), P30 AG062677 (PIs Brad Boeve, MD, Ronald Petersen, MD, PhD), P30 AG079280 (PI Jessica Langbaum, PhD), P30 AG062422 (PI Gil Rabinovici, MD), P30 AG066511 (PI Allan Levey, MD, PhD), P30 AG072946 (PI Linda Van Eldik, PhD), P30 AG062715 (PI Sanjay Asthana, MD, FRCP), P30 AG072973 (PI Russell Swerdlow, MD), P30 AG066506 (PIs Glenn Smith, PhD, ABPP, David Lowenstein, PhD, Ranjan Duara, MD), P30 AG066508 (PIs Stephen Strittmatter, MD, PhD, Christopher Van Dyck, MD), P30 AG066515 (PI Victor Henderson, MD, MS), P30 AG072947 (PI Suzanne Craft, PhD), P30 AG072931 (PI Henry Paulson, MD, PhD), P30 AG066546 (PIs Sudha Seshadri, MD, Gladys Maestre, MD, PhD), P30 AG086401 (PI Erik Roberson, MD, PhD), P30 AG086404 (PI Gary Rosenberg, MD), P30 AG086403 (PI Angela Jefferson, PhD), P30 AG072958 (PIs Heather Whitson, MD, Gwenn Garden, MD, PhD), P30 AG072959 (PI Jagan Pillai, MD, PhD), P30 AG092752 (Ihab Hajjar, MD, MS).

During the preparation of this manuscript, the authors used Claude Opus 5.5 (Anthropic) to support literature searching, code optimization and debugging, and language editing. All identified references were verified against the original publications, and the authors take full responsibility for the manuscript’s content.

