## Supplementary Material for "Diagnostic Utility of Multi-Domain Language Measures Across the Alzheimer Disease Continuum"

### Supplementary Materials

#### Supplementary Figure 1. Score Distributions and ROC Curves for Standard and Specialized Measures.

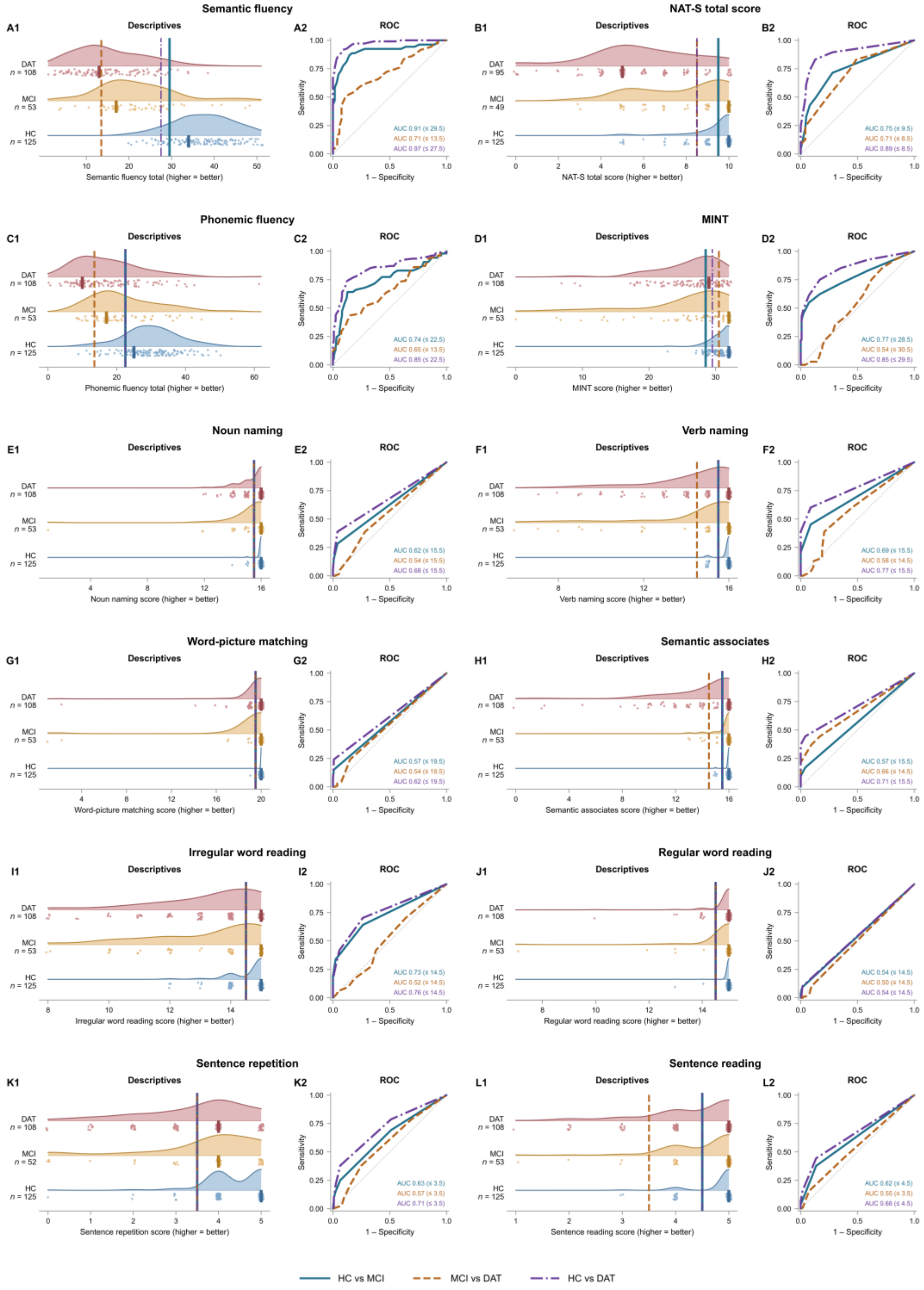

*Note*. **A1** and **A2** show semantic fluency; **B1** and **B2** show NAT-S total score; **C1** and **C2** show letter fluency; **D1** and **D2** show MINT; **E1** and **E2** show noun naming; **F1** and **F2** show verb naming; **G1** and **G2** show word-picture matching; **H1** and **H2** show semantic associates; **I1** and **I2** show irregular word reading; **J1** and **J2** show regular word reading; **K1** and **K2** show sentence repetition; **L1** and **L2** show sentence reading. **Panels A1–L1**: shaded curves are kernel density estimates, points are individual scores (jittered), and the vertical bar marks each group's modal score. Vertical reference lines mark the Youden J-optimal cut-off for each pairwise comparison, matched by colour and line style to the curves in Panels A2–L2. **Panels A2–L2**: the diagonal indicates chance performance. AUC and the corresponding Youden cut-off are printed within each ROC panel, colour-matched to the contrast. For NAT-S total score, the MCI–DAT and HC–DAT cut-offs coincide at 8.5. For letter fluency, the HC–MCI and HC–DAT cut-offs coincide at 22.5. For noun naming, the HC–MCI, MCI–DAT, and HC–DAT cut-offs coincide at 15.5. For verb naming, the HC–MCI and HC–DAT cut-offs coincide at 15.5. For word-picture matching, the HC–MCI, MCI–DAT, and HC–DAT cut-offs coincide at 19.5. For semantic associates, the HC–MCI and HC–DAT cut-offs coincide at 15.5. For irregular word reading, the HC–MCI, MCI–DAT, and HC–DAT cut-offs coincide at 14.5. For regular word reading, the HC–MCI, MCI–DAT, and HC–DAT cut-offs coincide at 14.5. For sentence repetition, the HC–MCI, MCI–DAT, and HC–DAT cut-offs coincide at 3.5. For sentence reading, the HC–MCI and HC–DAT cut-offs coincide at 4.5. AUC = area under the curve, with 95% bootstrap confidence intervals (2,000 resamples). MINT = Multilingual Naming Test; NAT-S total score = Northwestern Anagram Test – Short Form. HC = healthy control; MCI = mild cognitive impairment; DAT = Alzheimer's disease dementia.

**Supplementary Table 1. Diagnostic Performance of Standard and Specialized Language Measures Across Three Pairwise Contrasts.**

**A. HC vs. DAT**

| Measure | AUC [95% CI] | SE | Sens. [95% CI] | Spec. [95% CI] | Cutoff |
|---|---|---|---|---|---|
| **Standard tests (UDSv3)** | | | | | |
| Semantic fluency (animals and vegetables) | .973* [.955, .988] | .009 | .917 [.863, .964] | .912 [.860, .958] | ≤ 27.50 |
| Letter fluency (F and L) | .846* [.790, .896] | .027 | .741 [.657, .821] | .872 [.810, .927] | ≤ 22.50 |
| MINT | .854* [.805, .900] | .024 | .750 [.670, .826] | .832 [.769, .895] | ≤ 29.50 |
| **Specialized tests (FTLD module)** | | | | | |
| Regular word reading | .543 [.514, .577] | .016 | .102 [.049, .163] | .984 [.957, 1.000] | ≤ 14.50 |
| Irregular word reading | .760* [.702, .816] | .029 | .704 [.618, .786] | .736 [.658, .812] | ≤ 14.50 |
| Word-picture matching test | .617 [.576, .658] | .021 | .241 [.160, .324] | .992 [.974, 1.000] | ≤ 19.50 |
| Semantic associates test | .710* [.661, .760] | .025 | .444 [.355, .541] | .960 [.924, .992] | ≤ 15.50 |
| NAT-S: total score | .891* [.845, .935] | .023 | .832 [.755, .904] | .872 [.811, .928] | ≤ 8.50 |
| Sentence repetition test | .712* [.648, .775] | .032 | .380 [.287, .472] | .936 [.889, .976] | ≤ 3.50 |
| Noun naming | .676 [.628, .724] | .025 | .389 [.296, .476] | .960 [.924, .992] | ≤ 15.50 |
| Verb naming | .774* [.719, .827] | .027 | .602 [.505, .696] | .912 [.859, .959] | ≤ 15.50 |
| Sentence reading | .662 [.604, .716] | .028 | .444 [.346, .538] | .864 [.797, .923] | ≤ 4.50 |

**B. HC vs. MCI**

| Measure | AUC [95% CI] | SE | Sens. [95% CI] | Spec. [95% CI] | Cutoff |
|---|---|---|---|---|---|
| **Standard tests (UDSv3)** | | | | | |
| Semantic fluency (animals and vegetables) | .905* [.842, .962] | .031 | .868 [.774, .957] | .856 [.794, .915] | ≤ 29.50 |
| Letter fluency (F and L) | .745* [.653, .829] | .045 | .642 [.508, .761] | .872 [.811, .927] | ≤ 22.50 |
| MINT | .771* [.684, .854] | .043 | .547 [.412, .686] | .928 [.878, .969] | ≤ 28.50 |
| **Specialized tests (FTLD module)** | | | | | |
| Regular word reading | .540 [.501, .585] | .021 | .094 [.021, .182] | .984 [.959, 1.000] | ≤ 14.50 |
| Irregular word reading | .728* [.646, .807] | .042 | .642 [.509, .771] | .736 [.658, .811] | ≤ 14.50 |
| Word-picture matching test | .572 [.526, .623] | .026 | .151 [.060, .255] | .992 [.975, 1.000] | ≤ 19.50 |

| Measure | AUC [95% CI] | SE | Sens. [95% CI] | Spec. [95% CI] | Cutoff |
|---|---|---|---|---|---|
| Semantic associates test | .567 [.516, .625] | .028 | .170 [.075, .280] | .960 [.921, .992] | ≤ 15.50 |
| NAT-S: total score | .750* [.667, .827] | .041 | .714 [.575, .841] | .720 [.639, .795] | ≤ 9.50 |
| Sentence repetition test | .634 [.550, .718] | .044 | .250 [.130, .373] | .936 [.893, .976] | ≤ 3.50 |
| Noun naming | .624 [.556, .690] | .034 | .283 [.158, .417] | .960 [.923, .992] | ≤ 15.50 |
| Verb naming | .692 [.615, .764] | .038 | .453 [.314, .585] | .912 [.857, .960] | ≤ 15.50 |
| Sentence reading | .623 [.551, .696] | .037 | .377 [.250, .510] | .864 [.806, .919] | ≤ 4.50 |

**C. MCI vs. DAT**

| Measure | AUC [95% CI] | SE | Sens. [95% CI] | Spec. [95% CI] | Cutoff |
|---|---|---|---|---|---|
| **Standard tests (UDSv3)** | | | | | |
| Semantic fluency (animals and vegetables) | .710* [.625, .790] | .042 | .509 [.412, .607] | .887 [.794, .966] | ≤ 13.50 |
| Letter fluency (F and L) | .649 [.563, .733] | .043 | .435 [.343, .528] | .849 [.750, .943] | ≤ 13.50 |
| MINT | .540 [.441, .639] | .051 | .852 [.786, .916] | .283 [.164, .411] | ≤ 30.50 |
| **Specialized tests (FTLD module)** | | | | | |
| Regular word reading | .501 [.452, .547] | .025 | .102 [.049, .163] | .906 [.824, .980] | ≤ 14.50 |
| Irregular word reading | .515 [.421, .612] | .049 | .704 [.618, .788] | .358 [.232, .500] | ≤ 14.50 |
| Word-picture matching test | .539 [.468, .604] | .034 | .241 [.161, .321] | .849 [.745, .941] | ≤ 19.50 |
| Semantic associates test | .658 [.594, .720] | .033 | .370 [.283, .459] | .906 [.820, .977] | ≤ 14.50 |
| NAT-S: total score | .708* [.620, .792] | .045 | .832 [.755, .903] | .510 [.367, .649] | ≤ 8.50 |
| Sentence repetition test | .572 [.485, .659] | .045 | .380 [.290, .467] | .750 [.627, .865] | ≤ 3.50 |
| Noun naming | .545 [.473, .628] | .040 | .389 [.303, .485] | .717 [.591, .837] | ≤ 15.50 |
| Verb naming | .579 [.486, .676] | .047 | .389 [.301, .481] | .792 [.680, .898] | ≤ 14.50 |
| Sentence reading | .546 [.464, .626] | .042 | .157 [.094, .230] | .925 [.844, .983] | ≤ 3.50 |

*Note.* * = AUC ≥ .700. Sens. = sensitivity. Spec. = specificity. Optimal cutoff determined by Youden's index. 95% CIs for sensitivity and specificity computed via bootstrap (2,000 replications). MINT = Multilingual Naming Test. NAT-S = Northwestern Anagram Test—Short Form.

**Supplementary Table 2. Pairwise Comparisons of Areas Under the ROC Curve (DeLong's Test) Against the Best-Performing Standard and Specialized Measures.**

**A. Reference: Semantic fluency (best standard measure)**

| Comparison measure | DAT vs. HC | | | MCI vs. HC | | | DAT vs. MCI | | |
|---|---|---|---|---|---|---|---|---|---|
| | ΔAUC | *z* | *p* | ΔAUC | *z* | *p* | ΔAUC | *z* | *p* |
| Regular word reading | .430 | 25.12 | < .001 | .366 | 10.64 | < .001 | .209 | 4.87 | < .001 |
| Word-picture matching | .357 | 16.39 | < .001 | .333 | 8.94 | < .001 | .171 | 3.94 | .008 |
| Sentence reading | .311 | 11.09 | < .001 | .282 | 6.80 | < .001 | .165 | 3.69 | .022 |
| Noun naming | .297 | 12.13 | < .001 | .282 | 7.19 | < .001 | .165 | 4.04 | .006 |
| Semantic associates | .264 | 10.59 | < .001 | .338 | 8.80 | < .001 | .052 | 1.22 | 1.000 |
| Sentence repetition | .261 | 8.35 | < .001 | .269 | 5.61 | < .001 | .140 | 2.69 | .625 |
| Irregular word reading | .213 | 7.55 | < .001 | .177 | 4.26 | .002 | .195 | 4.26 | .002 |
| Verb naming | .199 | 7.83 | < .001 | .214 | 5.36 | < .001 | .131 | 2.91 | .331 |
| Letter fluency | .128 | 5.36 | < .001 | .160 | 4.10 | .003 | .062 | 1.64 | 1.000 |
| MINT | .119 | 5.04 | < .001 | .134 | 3.17 | .112 | .170 | 3.83 | .013 |
| NAT-S total | .079 | 3.58 | .017 | .147 | 3.14 | .119 | .012 | 0.22 | 1.000 |

**B. Reference: NAT-S total score (best specialized measure)**

| Comparison measure | DAT vs. HC | | | MCI vs. HC | | | DAT vs. MCI | | |
|---|---|---|---|---|---|---|---|---|---|
| | ΔAUC | *z* | *p* | ΔAUC | *z* | *p* | ΔAUC | *z* | *p* |
| Regular word reading | .342 | 13.20 | < .001 | .207 | 4.96 | < .001 | .205 | 4.47 | < .001 |
| Word-picture matching | .274 | 8.44 | < .001 | .183 | 4.25 | .002 | .163 | 2.94 | .299 |
| Sentence reading | .236 | 7.29 | < .001 | .132 | 2.37 | .965 | .165 | 3.03 | .227 |
| Noun naming | .204 | 6.63 | < .001 | .136 | 2.81 | .325 | .146 | 2.52 | .995 |
| Semantic associates | .189 | 6.46 | < .001 | .187 | 4.29 | .002 | .053 | 1.16 | 1.000 |
| Sentence repetition | .176 | 4.82 | < .001 | .108 | 1.92 | 1.000 | .129 | 2.32 | 1.000 |
| Verb naming | .119 | 4.10 | .002 | .083 | 1.74 | 1.000 | .104 | 2.03 | 1.000 |
| Irregular word reading | .114 | 3.40 | .033 | .025 | 0.53 | 1.000 | .181 | 3.35 | .078 |
| Semantic fluency | −.079 | −3.58 | .017 | −.147 | −3.14 | .119 | −.012 | −0.22 | 1.000 |
| Letter fluency | .058 | 1.91 | 1.000 | .007 | 0.12 | 1.000 | .064 | 1.18 | 1.000 |
| MINT | .026 | 0.84 | 1.000 | −.012 | −0.24 | 1.000 | .144 | 2.48 | 1.000 |

*Note.* Each row compares the listed measure against the reference measure for that panel. ΔAUC = AUC(reference) − AUC(comparison measure), so positive values indicate the reference discriminated better; *z* and *p* are from DeLong's test for two correlated ROC curves (DeLong et al., 1988). NAT-S = Northwestern Anagram Test, sentence subtest; MINT = Multilingual Naming Test; HC = healthy control; MCI = mild cognitive impairment; DAT = Dementia of Alzheimer's type. *p* values are Holm–Bonferroni corrected; $^{*} = p < .05$. $^{**} = p < .01$. $^{***} = p < .001$.

**Supplementary Table 3. Generalized Variance Inflation Factors for the Hierarchical Regression Model.**

| Predictor | df | GVIF | Adj. GVIF |
|---|---|---|---|
| Age | 1 | 1.09 | 1.04 |
| Sex | 1 | 1.03 | 1.02 |
| Education | 1 | 1.03 | 1.01 |
| Diagnosis | 2 | 2.70 | 1.29 |
| TMT-B | 1 | 2.89 | 1.70 |
| Number Span Test: Backward | 1 | 1.70 | 1.30 |

*Note*. GVIF = generalized variance inflation factor. Adj. GVIF = GVIF^(1/(2·df)), the degrees-of-freedom–adjusted value comparable across predictors with differing degree of freedom. All adjusted values are below 2, indicating no problematic multicollinearity. TMT-B = Trail Making Test Part B.

**Supplementary Table 4. Hierarchical Regression Model Summary Predicting NAT-S Total Score.**

| Step | Block | $R^2$ | Adj. $R^2$ | $\Delta R^2$ | $\Delta F$ | $df_1$ | $df_2$ | p |
|---|---|---|---|---|---|---|---|---|
| 1 | Age, sex, education, diagnosis | .261 | .243 | .261 | 14.81 | 5 | 210 | < .001 |
| 2 | + TMT-B, Number Span Backward | .357 | .335 | .096 | 15.56 | 2 | 208 | < .001 |

*Note.* TMT-B = Trail Making Test Part B. $\Delta R^2$ = change in $R^2$ at each step; $\Delta F$ = F-change statistic.

**Supplementary Table 5. Regression Coefficients Predicting NAT-S Total Score.**

| Step | Predictor | B | SE | 95% CI | t | p | $sr^2$ |
|---|---|---|---|---|---|---|---|
| 1 | Intercept | 8.416 | 1.270 | [5.913, 10.919] | 6.63 | < .001 | |
| 1 | Age | .009 | .016 | [−.022, .040] | 0.56 | .576 | .0011 |
| 1 | Sex | .372 | .232 | [−.085, .829] | 1.60 | .110 | .0091 |
| 1 | Education | −.006 | .051 | [−.107, .095] | −0.12 | .905 | .0001 |
| 1 | Diagnosis: MCI | −1.633 | .304 | [−2.233, −1.033] | −5.37 | < .001 | .1014 |
| 1 | Diagnosis: DAT | −2.187 | .290 | [−2.759, −1.615] | −7.54 | < .001 | .2002 |
| 2 | Intercept | 8.133 | 1.293 | [5.583, 10.683] | 6.29 | < .001 | |
| 2 | Age | .013 | .015 | [−.017, .042] | 0.85 | .394 | . 0023 |
| 2 | Sex | .273 | .218 | [−.158, .703] | 1.25 | .213 | .0048 |
| 2 | Education | −.017 | .048 | [−.112, .078] | −0.35 | .724 | .0004 |
| 2 | Diagnosis: MCI | −.727 | .329 | [−1.376, −.079] | −2.21 | .028 | .0151 |
| 2 | Diagnosis: DAT | −.581 | .410 | [−1.389, .226] | −1.42 | .157 | .0062 |
| 2 | TMT-B | −.008 | .002 | [−.012, −.004] | −3.81 | < .001 | .0448 |
| 2 | Number Span Test: Backward | .116 | .055 | [.008, .224] | 2.11 | .036 | .0138 |

*Note.* Diagnosis is dummy-coded with healthy controls as the reference group. *B* = unstandardized coefficient; *SE* = standard error; $sr^2$ = squared semi-partial correlation. HC = healthy controls; MCI = mild cognitive impairment; DAT = Dementia of Alzheimer's type. TMT-B = Trail Making Test Part B.
